# Novel meta-analysis pipeline of heterogeneous high-throughput gene expression datasets reveals dysregulated interactions and pathways in asthma

**DOI:** 10.1101/19012377

**Authors:** Brandon Guo, Abhinav Kaushik, Kari C. Nadeau

## Abstract

**Introduction:** Asthma is a complex and chronic inflammatory disorder with varying degrees of airway inflammation. It affects ∼235 million people worldwide, and about 8% of the United States population. Unlike single-gene disorders, asthma phenotypes are guided by a highly variable combination of genotypes, making it a complex disease to study computationally. Recently, several independent high-throughput gene expression studies in bioinformatics have identified and proposed numerous molecular drivers involved in asthma initiation and progression. However, there is a poor consensus in our understanding of the molecular factors involved in the mechanism of this disease due to inherent genetic heterogeneity. Such an uncertainty in bioinformatics studies have led to a “reproducibility crisis” in the field, where similar analyses can often yield greatly varying results. In this study, we seek to harness heterogeneity in asthma by applying a meta-analysis that explores varying tissue environments.

**Methods:** We use three publicly-available microarray gene expression datasets, belonging to different tissues in asthma patients, from NCBI’s Gene Expression Omnibus (GEO). As a meta-analysis, we apply a mixed-model effect size test to determine differentially expressed (DE) genes across all three studies. Then, The datasets are pre-processed and subjected to Weighted Gene Co-expression Network Analysis (WGCNA) for identification of functional modules. Using module preservation, we determine modules in asthma that were not preserved in the healthy condition, then combine the three with a Fisher’s test for a set of asthma-unique modules. These modules are explored using functional analysis (i.e. GO term analysis). Using the DE genes as well as known transcription factors, we re-construct Gene Regulatory Networks (GRNs) for each of our shortlisted modules. We then studied the topology of these GRNs using hive plots to reveal underlying dysregulations, paving the way for future analyses.

**Results:** Our analysis reveals a novel perspective to a key interaction in asthma inflammatory regulation, the CHD4-CCL26 transcription relation. Our hive plot analysis is able to explore this gene interaction beyond the typical “over-expression, under-expression” results from typical bioinformatics studies. We reveal that CCL26, an important regulator of asthma, appears to increase in expression and topological degree in asthma, but loses connection to CHD4, which seems to be characteristic to the asthma disease. Such analysis suggests that the topology of gene networks, above simply expression values, may be key to understanding the nuanced interactions between fundamental biomarkers and drug targets in complex diseases like asthma.

## Introduction

Asthma is a complex type I hypersensitivity and chronic inflammatory disorder with varying degrees of airway inflammation ^1^. It affects ∼358 million people worldwide and its origin constitutes overlapping environmental, genetic, and behavioral factors ^1,2^. Often, it co-exists as a comorbidity and shows large heterogeneity in response to available asthma medications ^3^. In the last decade, genotyping technologies, e.g. microarray and RNA-seq, have elucidated a variety of dysregulated genetic components that are associated with asthma initiation and progression ^4,5^. Despite the fact that asthma is a heterogeneous disorder, the identification of reliable disease drivers still remains a fundamental challenge ^6^.

Although gene expression-based studies have improved our understanding about the genetic perturbation associated with asthma, the findings are strongly affected by the disease heterogeneity and sample-specific variation, making the result inconsistent across independent studies ^7–9^. To overcome the problem, meta-analysis of multiple studies has been used multiple times to report common disease drivers across independent datasets ^1,8,10^. The rationale behind these *meta-analyses* is that the investigation of individual-cohort of subjects may not achieve genome/transcriptome-wide statistical significance; however, collaboration between multiple independent studies can increase the sample size and statistical power ^1,11^. Another approach to address the problem of heterogeneity involves the study of the interaction among groups of genes, i.e. modules, which are investigated at the systems-level to identify genes associated with common function in a given biological state ^12,13^. Here the rationale is that a strongly co-expressed genes group are likely to have more association and impact on a given phenotypic state than individual genes ^14–16^. In addition, these pathway-based approaches do not rely on a specific set of genes and larger, more tangible biological processes and pathways can be studied with higher reproducibility and accuracy than specific genes ^5,16^.

In asthma, structural remodeling of the large and small airways has been documented in all degrees of asthma severity ^17^. The airway remodeling has been linked to chronic inflammatory process, largely controlled by environmental factors, and involves activation of inflammatory cells including CD4+ T cells, mast cells and neutrophils ^6,18^. Evidence also suggests the role of genetic perturbations in declining the lung function during the remodeling process ^2^. Previously, meta-analysis of gene expression in airway epithelium of asthma identified several dysregulated genes and associated pathway based on 355 cases and 193 controls samples ^19^. Their findings reveal 1273 differentially expressed genes of which ∼35% have been found to be consensus irrespective of airway epithelium (bronchial and nasal) and demographic factors ^19^. Overall, the analysis from independent studies suggest that the airway epithelium plays a crucial role in disease pathogenesis and provide information about the exacerbations and therapeutic response ^20,21^. Despite the importance in understanding asthma pathogenesis, there are limited number of meta-analysis studies that attempts to elucidate the conserved elements associated with transcriptomic changes across different regions of airway epithelium.

Therefore, in this study, we are interested in studying the airway epithelium as a lens into the asthma disease. We apply a unique meta-analysis approach on publicly available gene expression datasets to elucidate the components conserved across transcriptomic profiles of different epithelial tissues of asthmatic patients. Our pipeline involves analysis of system-level (network module-based), gene-level, and gene regulatory-level information to generate consensus genes and pathways that are reproducibly affected in asthma across different airway epithelium in independent datasets.

## Results

### Data pre-processing

Three sets of microarray data were downloaded from the Gene Expression Omnibus (GEO) and formed into matrices with gene IDs as the row names and patient identifiers as the column names. In Table 1, we present basic information about our microarray matrices and their authors, for future reference. Multiple probe readings corresponding to one gene identifier are combined by averaging and probe readings with no identifiers were removed. Quantile normalization is performed on each of the three matrices. Boxplots for each study are plotted in Figure S1, with the author names used to identify the datasets. The normalized and pruned matrices are available for download in Supplementary File 1.

**Table 1.** Overview of public microarray datasets used in this paper. Individual studies will be referred to by author name as identification.

| **GEO ID** | **Epithelia Source** | **Healthy Patients** | **Asthma Patients** | **Reference** |
| --- | --- | --- | --- | --- |
| **GSE64913** | Bronchial | 23 | 13 | *Singhania et al.* |
| **GSE63142** | Bronchial | 27 | 125 | *Wenzel et al.* |
| **GSE65204** | Nasal | 33 | 36 | *Yang et al.* |

### Gene-expression meta-analysis

The publicly available microarray datasets belonging to three different airway epithelia were obtained from Gene Expression Omnibus (GEO; Table 1). The quantile normalized gene expression dataset (Figure S1) quality was evaluated by analyzing the conserved differentially expressed (DE) genes across different regions of airway epithelium using MetaIntegrator ^8^. We select GSE64913 and GSE65204 for our meta-analysis and use GSE63142 as a validation cohort.

The meta-analysis reveals 1,011 genes found to be differentially expressed (DE; effect size test with p-value < 0.1) across healthy and asthmatic patients across all the two training-set tissues. In Figure 1, we present a ROC curve of our validation cohort with respect to the 1,011 DE genes. The results are in agreement with previous findings that suggest the presence of common aberrations in the gene expression profiles of asthmatic patients, irrespective of airway epithelium ^19,22^. To further highlight the differential nature of these genes, we present in Figure 2 a violin plot of the effect size values in the independent cohort (GSE63142) of the genes we selected. The p-value is presented to show the differential expression in the independent study.

**Figure 1.**
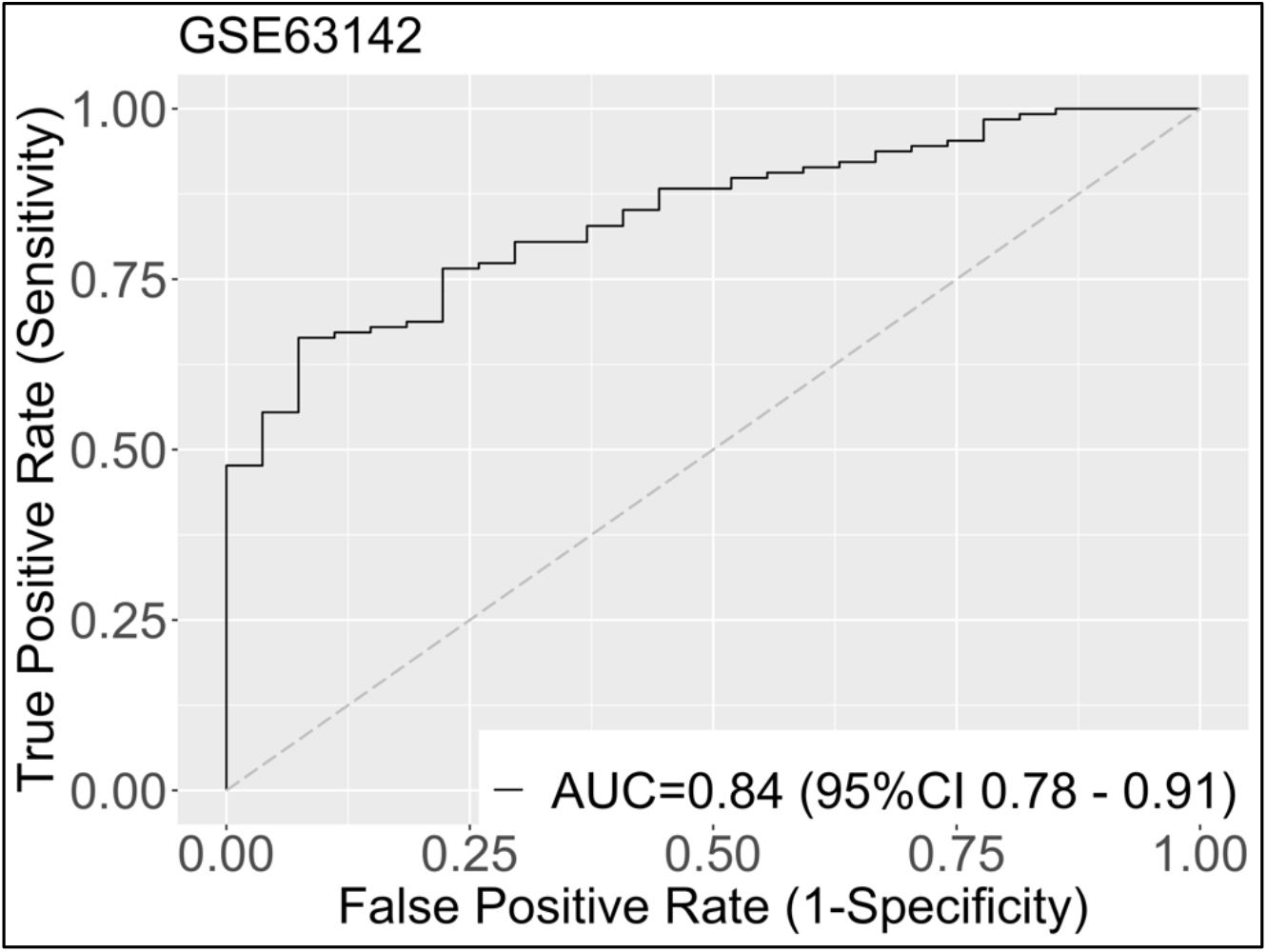
Receiver operating characteristic (ROC) curve of classification using bronchial and nasal microarray data. The AUC score of 0.84 suggests that different epithelial sources have common underlying genetic players.

**Figure 2.**
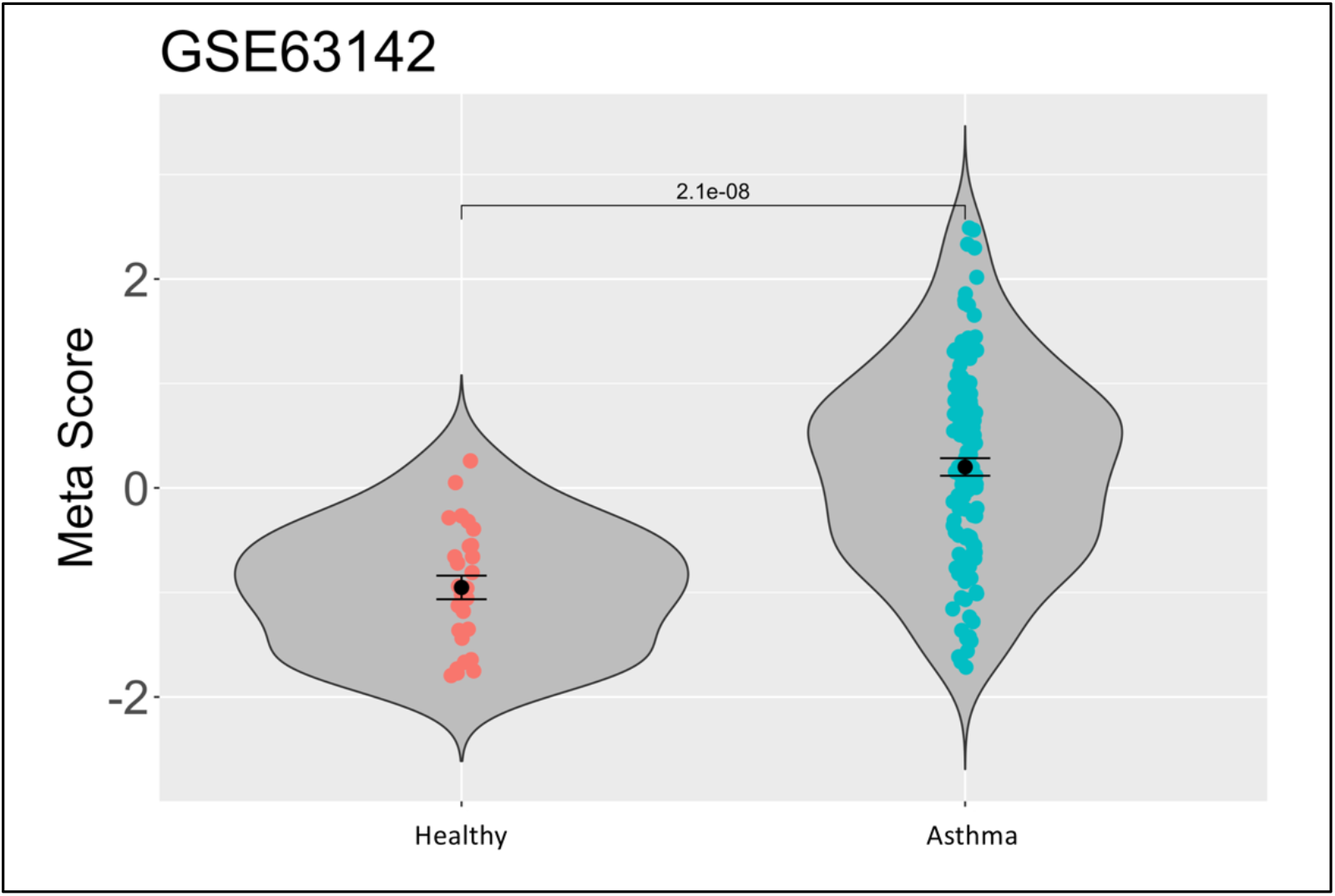
Violin plot of effect size scores in both conditions suggests that there are fundamental differences in effect size across asthma and healthy conditions, with significant p-value.

Among the 1200 genes, a large proportion of genes are already known to play active role in asthma initiation and progression. For example, one of the most under-expressed genes in asthma is the CD74 gene, and the most over-expressed gene is the CD44 gene. In Figure 3, we present the forest plots of their effect size expression across our two heterogeneous datasets. There is strong evidence in asthma literature that the CD44 and CD74 genes are key to inflammatory pathways that show dysregulation in asthma ^23–25^.

**Figure 3.**
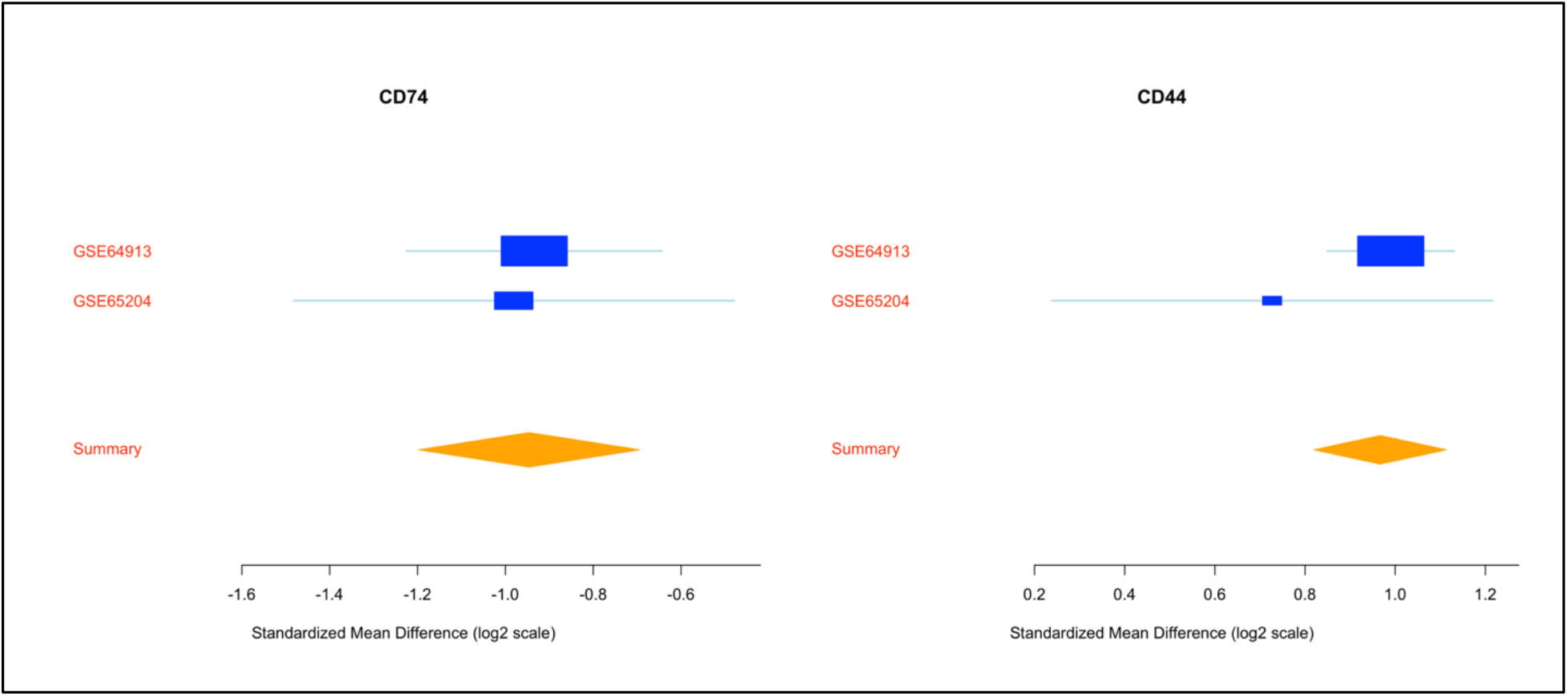
Forest plot of CD74, a heavily under-expressed gene in asthma, and CD44, a heavily over-expressed gene in asthma. Both genes are well-established to be biomarkers of asthma.

The complete list of DE genes, statistical significance and the associated functional role is available in supplementary file S2. We perform functional analysis on the entire list of the DE genes using the TargetMine database ^26^. Table 2 reveals key pathways that are enriched by these differentially expressed genes and their statistical significance as given by the Benjamini-Hochberg p-value correction method ^27^. The analysis reveals that the broad theme of dysregulated pathways, across different tissues, consists of those related to cell differentiation in Th-cells, which is again in agreement with the previous findings ^4,28^.

**Table 2.** Key asthma-related pathways show significant overlap, as given by the TargetMine database, with the differentially expressed genes found by MetaIntegrator.

| Pathway name | p-value (BH corrected) |
| --- | --- |
| Antigen processing and presentation | 2.88e-11 |
| Th1 and Th2 cell differentiation | 2.03e-8 |
| Asthma | 5.18e-7 |
| Th17 cell differentiation | 8.56e-7 |

### Gene co-expression network reconstruction

In order to investigate the conserved changes in the molecular interactions, we performed the Weighted Gene Co-expression Network Analysis (WGCNA) on the gene expression datasets to reconstruct six independent co-expression networks (see materials and methods). Macro-level properties of these six networks can be found in Table 3.

**Table 3.**
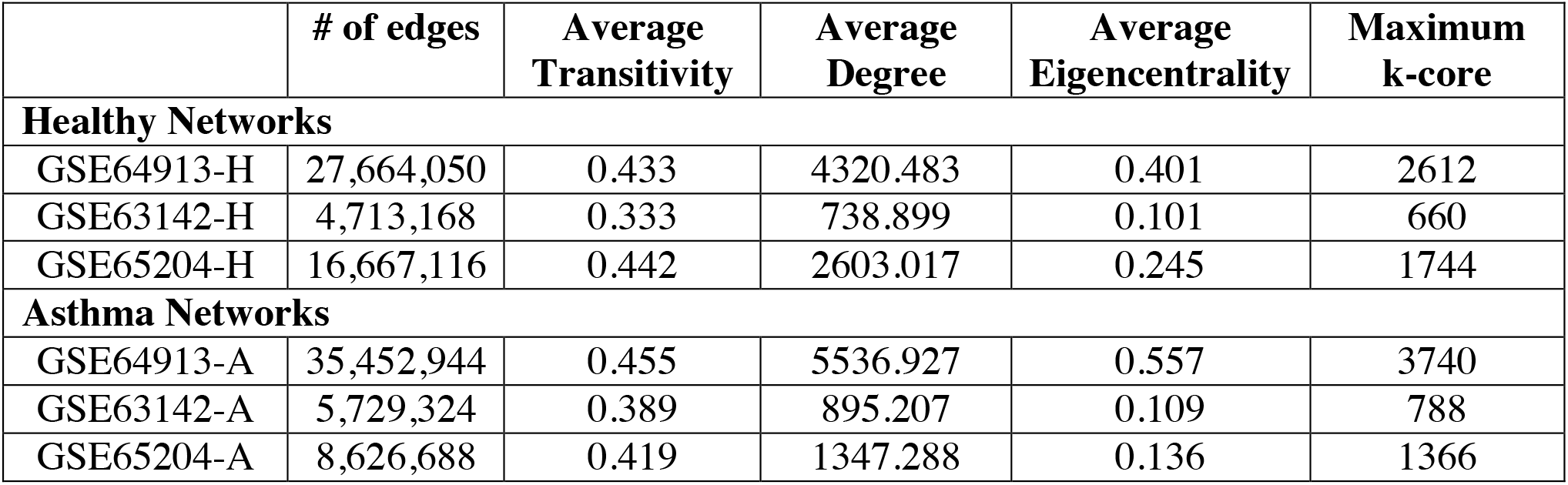
Network topology properties of the WGCNA networks for edges above cutoff of r > 0.1.

|  | # of edges | Average Transitivity | Average Degree | Average Eigencentrality | Maximum k-core |
| --- | --- | --- | --- | --- | --- |
| **Healthy Networks** |  |  |  |  |  |
| GSE64913-H | 27,664,050 | 0.433 | 4320.483 | 0.401 | 2612 |
| GSE63142-H | 4,713,168 | 0.333 | 738.899 | 0.101 | 660 |
| GSE65204-H | 16,667,116 | 0.442 | 2603.017 | 0.245 | 1744 |
| **Asthma Networks** |  |  |  |  |  |
| GSE64913-A | 35,452,944 | 0.455 | 5536.927 | 0.557 | 3740 |
| GSE63142-A | 5,729,324 | 0.389 | 895.207 | 0.109 | 788 |
| GSE65204-A | 8,626,688 | 0.419 | 1347.288 | 0.136 | 1366 |

**Table 3.** Overview of module counts in each of the studies and number of dysregulated modules in asthma as given by the Zsummary permutation test (WGCNA).

|  | *Singhania et al.* | *Wenzel et al.* | *Yang et al.* |
| --- | --- | --- | --- |
| # Healthy modules | 33 | 55 | 30 |
| # Asthma modules | 42 | 82 | 38 |
| # Dysreg. modules<br>(Zsummary < 10) | 31 | 27 | 8 |

Subsequently, we initiate the network analysis by evaluating the scale-free topologies in all of the resultant networks. The networks associated with each power are compared to a scale free network, and correlation coefficient is computed. We are interested in those powered networks who mimic scale-free topology closely. We choose a given value of soft threshold based on scale-free topology; this selection is shown in Figure 3.

Our WGCNA networks now follow scale-free thresholds as shown by the log-log distributions of degree densities of the networks presented in Figure 4: a downward linear trend implies that the data exhibits scale-free topology. Scale free topology allows us to highlight hub genes and key interactions ^29,30^.

**Figure 4.**
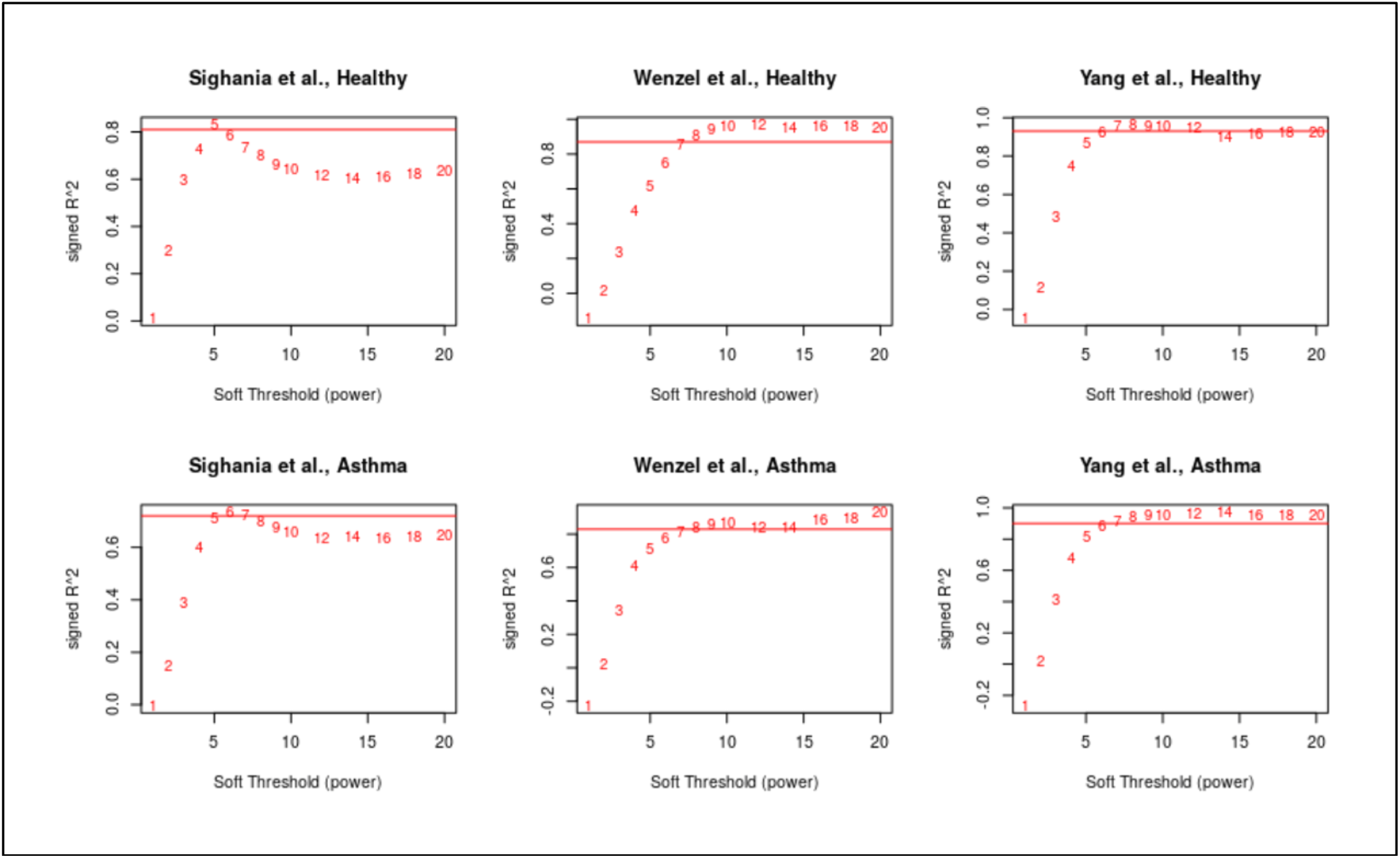
Correlation to scale-free topology based on power of soft-thresholding. Pearson’s coefficient is used to select power for resulting, adjusted WGCNA networks.

The analysis of transitivity distributions in Figure 5 suggests the elevated loss of gene-gene connections in the asthmatic condition evident with the decrease in the number of genes with high transitivity. Loss of gene connections during the disease state has been observed in a number of previous studies and it has been found to be linked with functional significance to disrupts normal cell robustness and drives disease progression ^6,31^.

**Figure 5.**
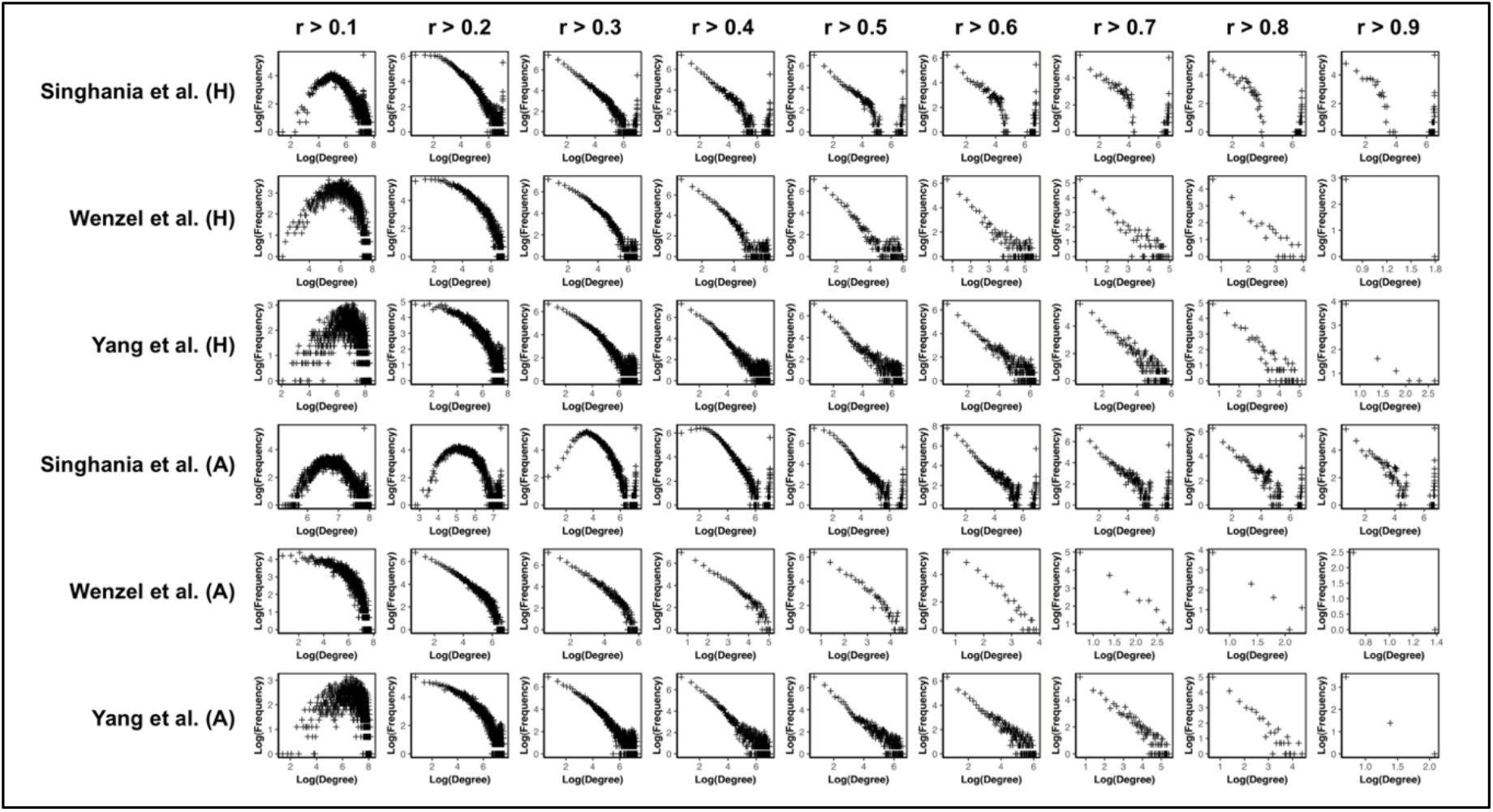
Log-log plots of degree and frequency based on different thresholds for the WGCNA networks after soft-thresholding. Negative linear relationship, especially at high cutoffs, suggests these networks closely resemble scale-free topology, allowing for identification of hub genes.

### Module preservation in networks

After establishing the six networks that possess the key scale-free topology, we seek to reduce the networks using modules, which represent biological function, from different studies into a coherent image to compare across asthma and healthy conditions.

To establish the modules that make up each network, we use the WGCNA clustering algorithm that uses a hierarchical clustering, dynamic tree-cut method to determine clusters. In Figure 6, we show dendrograms for each of the three healthy networks. Using a tree-cut threshold of 0.99, we establish the modules shown in the bars below. The two bars are colored based on how a certain sequence (the one given by the WGCNA cluster on healthy) of genes is clustered. Comparing the two color distributions, it appears that there is low module preservation across healthy and asthma in these studies. Concretely, genes that tend to cluster in healthy do not show the same behavior in asthma. This is further evidence of the genome-wide dysregulation that occurs in asthma.

**Figure 6.**
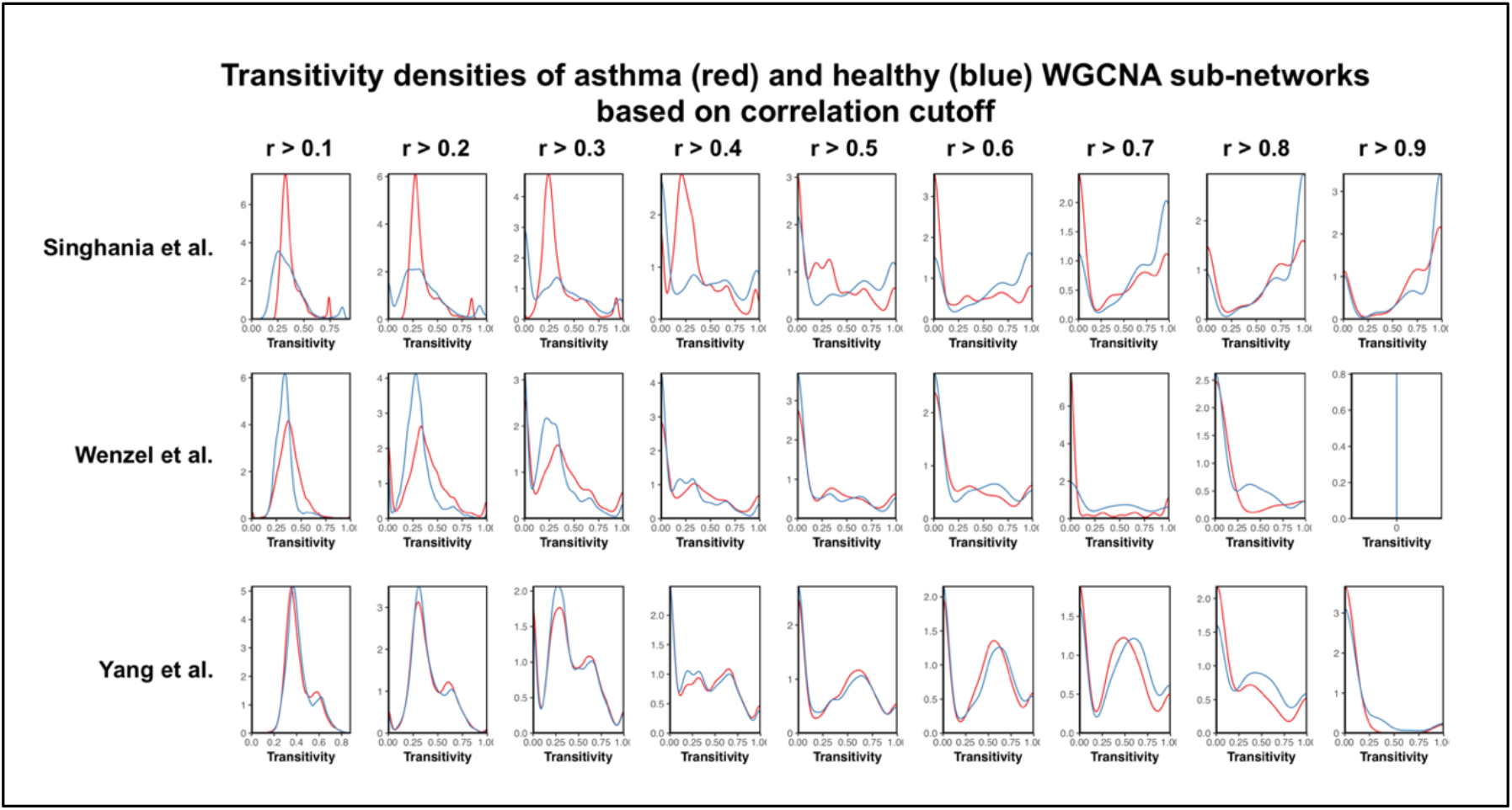
Transitivity density plots for WGCNA networks. At high transitivity values, asthma distribution seems to be more sparse, suggesting a general dysregulation of gene interactions.

We are interested in determining the modules of genes in asthma that show little preservation in healthy. To achieve this, we determine the Zsummary score for each module in asthma, which represents how much a given module in asthma clusters in healthy. All modules that have Zsummary scores under 10 are determined to not be significantly preserved across conditions. We extract all such modules as ones that may be dysregulated in asthma.

Table 3 shows a summary of the properties of our modules and networks. It indicates how many modules we eventually extract from each network for having a Zsummary score below 10.

### Gene prioritization

Of the modules in asthma that we have identified as those that do not retain structural preservation in the healthy condition, we identify the key genes using gene ontology (GO) terms and the parent terms associated with their hierarchies.

For each of the dysregulated modules, we determine at a threshold of p = 0.001 the GO terms that are highlighted by the genes in each module. Based on the GO term hierarchy, we find one parent term, which is the highlighted term most representative of the gene sample, to correspond with one module. In Tables 4-1, 4-2, and 4-3, we highlight GO terms that are parent GO terms of more than one module in the network. Associated with these GO terms are the hub genes of each module that have this parent GO term. These hub genes are those that are filtered to be correlated with functional-level dysregulations in asthma.

**Table 4-1.** Hub genes associated with highlighted parent GO terms in Singhania et al.

| **Parent GO term** | **Healthy Hub Genes** | **Asthma Hub Genes** |
| --- | --- | --- |
| reproduction | CDO1, CST9L, NAT2 | CMA1, PRAC1 |
| negative regulation of RNA transcription | N/A | NCR1, C1orf61 |
| MAPK cascade | HOXA13, SYT16, CDC45 | CD81-AS1, RGS9 |
| nucleotide binding | VENTX, CLC | N/A |
| lytic vacuole | CNDP1, BRDT | N/A |
| transcription regulatory region, DNA binding | N/A | PNMA6A, IFNA14, PLAC1 |
| cell activation | N/A | MYH4, CALB2, IL25 |
| molecular function | N/A | LMOD1, GPR19 |

**Table 4-2.**
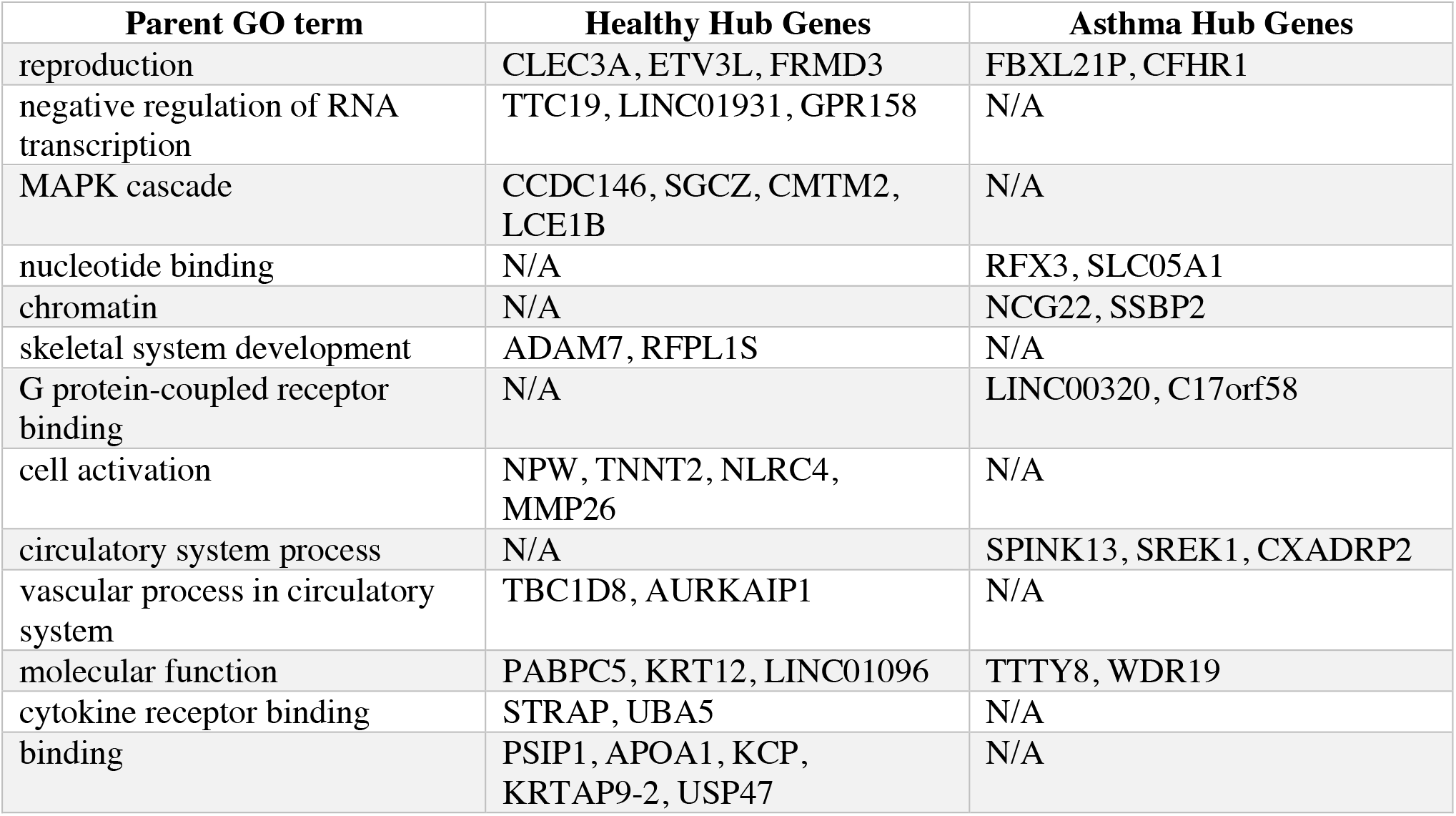
Hub genes associated with highlighted parent GO terms in Wenzel et al.

**Table 4-3.**
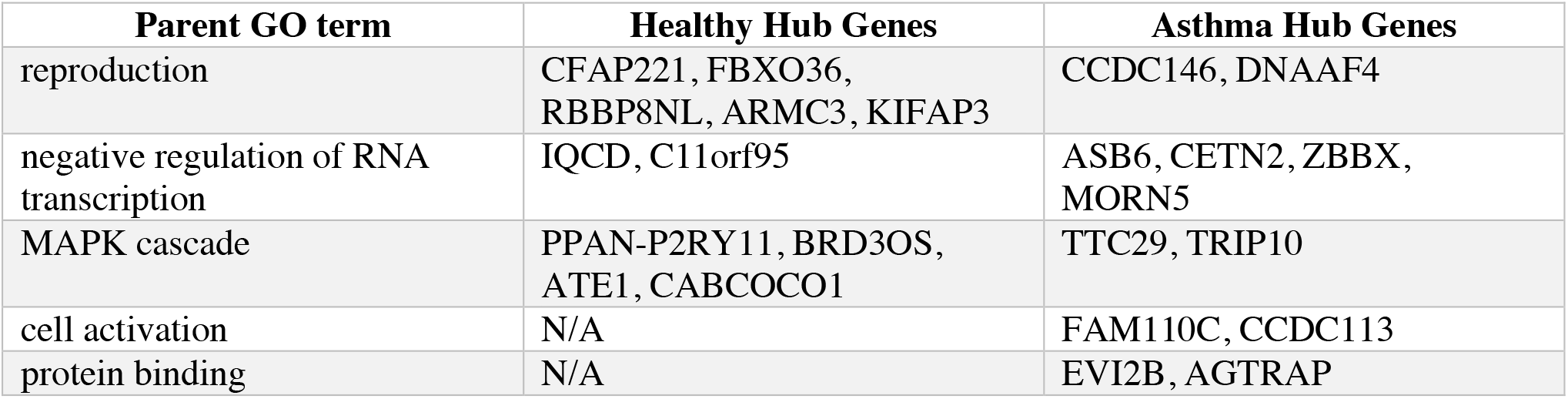
Hub genes associated with highlighted parent GO terms in Yang et al.

### Identification of asthma-unique modules

We define an “asthma-unique” module as a module in asthma that consistently shows structural and functional dysregulation when compared to modules in the associated healthy condition. To identify these modules, which is a strict subset of the weakly preserved modules previously identified, we perform a Fisher’s exact test across the three sets of networks.

Figure 8 shows a heatmap that compares the unpreserved modules (labelled A, B, C based on the study they come from) with the unpreserved modules from the other two studies. The color of each individual element of the heatmap represents its p-value with another module. We establish a threshold for the Fisher’s test for overlap of p < 0.05. We are interested in all modules that share a significant similarity value with at least one other module in *both* other networks.

**Figure 7.**
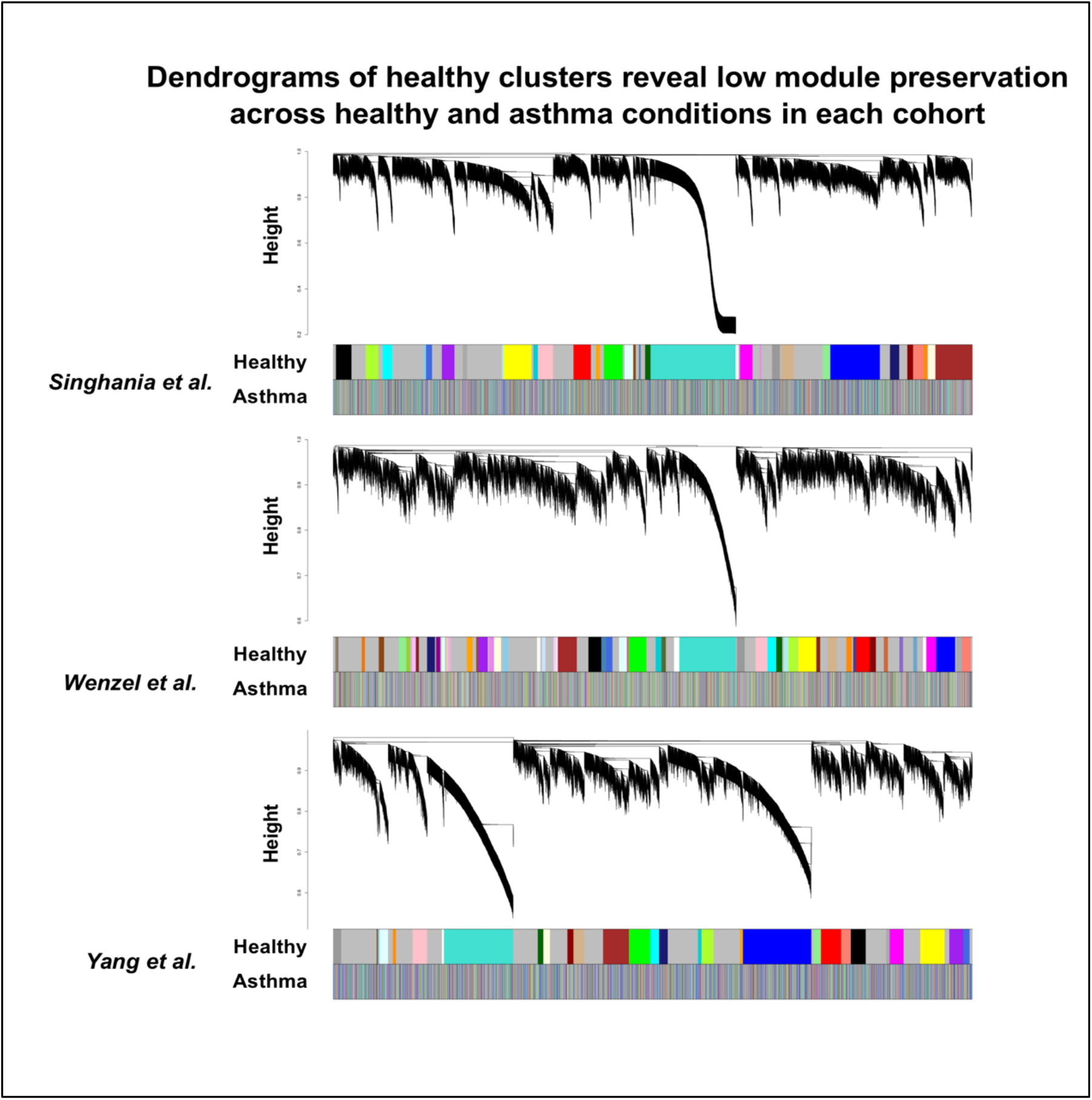
Dendrograms of clustering in healthy condition, with asthma clusters below. Colors of the “Healthy” and “Asthma” bars represent the modules of the genes with respect to their horizontal position in the dendrogram. Bars show generally low module preservation in asthma.

**Figure 8.**
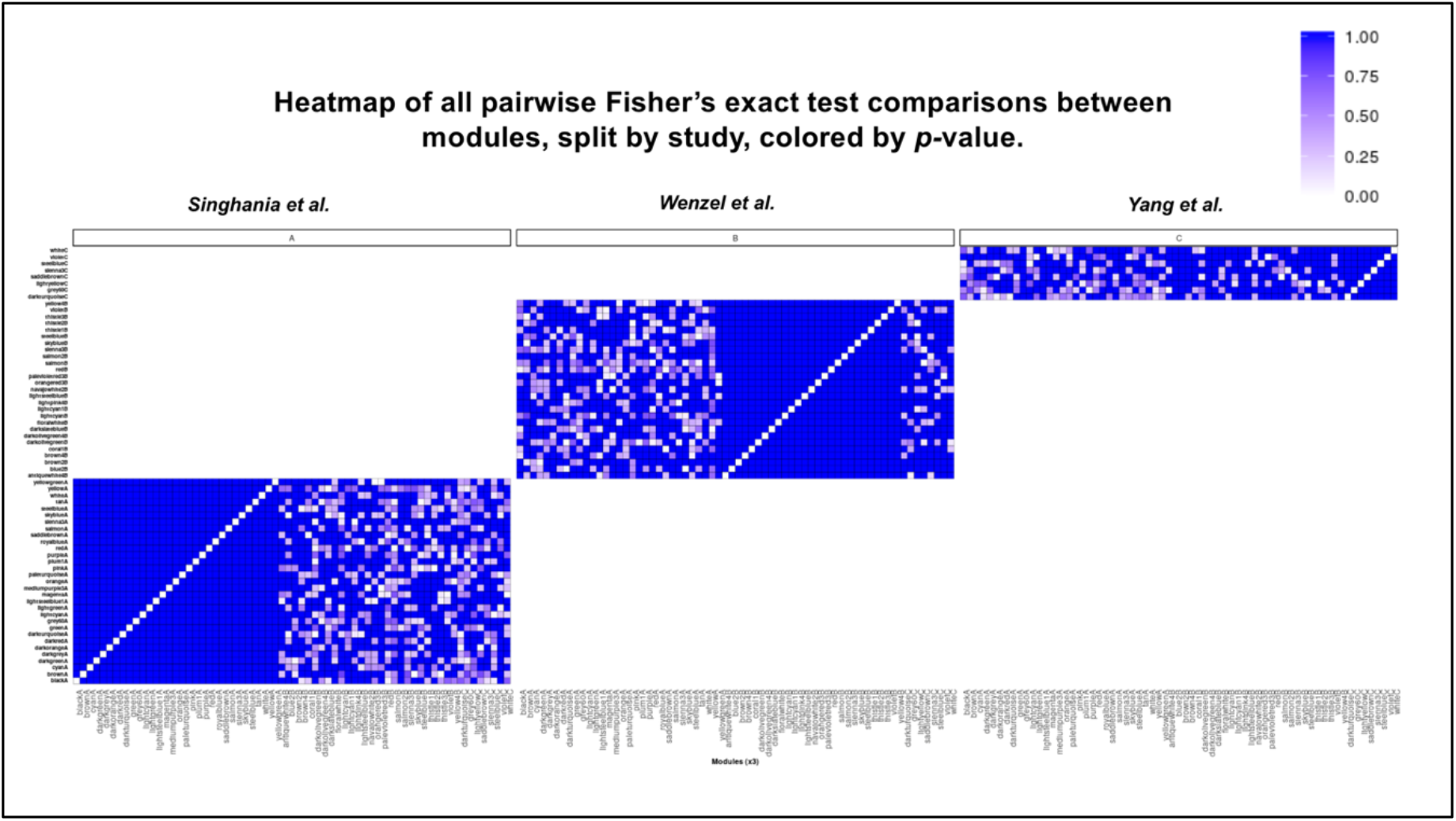
Heatmap of p-values of Fisher’s exact test across the dysregulated modules in asthma.

Figure 9 shows the same heatmap in Figure 8, but only p-values of 0.05 or less are distinguished by the white color. We also highlight the modules that satisfy the condition of having conservation in both other networks. These are all the “asthma-unique” modules.

**Figure 9.**
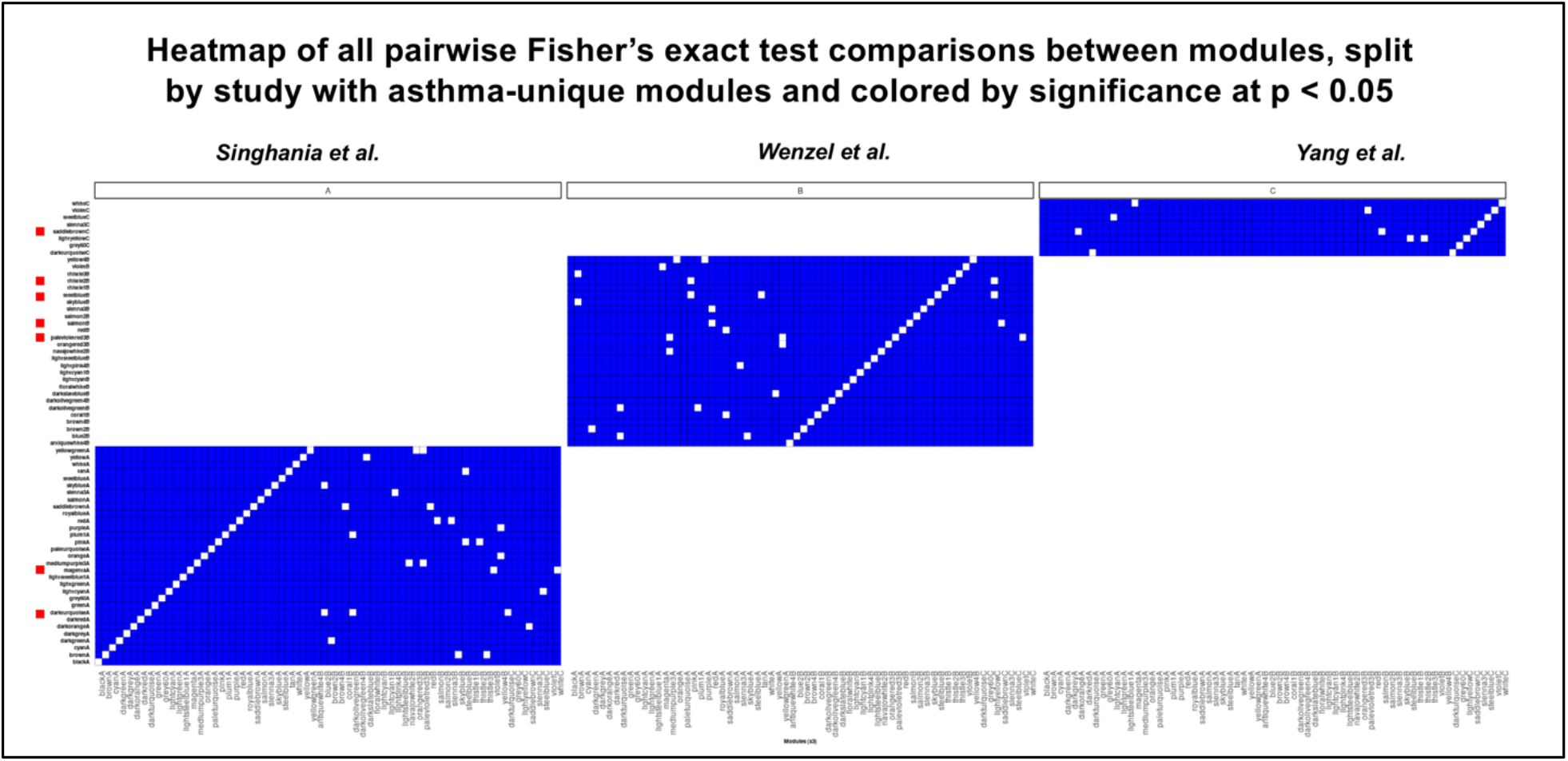
Heatmap of significance of similarity across dysregulated modules in asthma. Those modules (highlighted in red) with significant similarity with a module in both other studies is considered “asthma-unique.”

In Table 5, we present all “asthma-unique” modules as well as GO term analysis of their gene sets. Because these are modules that we hypothesize play a functional role in their own networks, we expect a generally consistent series of highly significant GO terms.

**Table 5.** GO term analysis of “asthma-unique” modules found in the three studies. “MagentaA” module is selected based on its critical role in asthma disease yet consistent dysregulation.

| **Module** | **Size** | **GO Terms** |
| --- | --- | --- |
| **darkturquoiseA** | 158 | protein transport, peptide transport, amide transport, protein localization, nitrogen compound transport, digestive tract development, |
| **magentaA** | 274 | cellular component organization, regulation of cellular process/biological process, signaling, cell communication, developmental process. |
| **palevioletred3B** | 46 | anatomical structure, organ morphogenesis, regulation of cellular process/glutamate secretion. |
| **salmonB** | 120 | thrombopoietin-mediated signaling, regulation of biological process, cellular process, regulation of biological quality, homeostatic process, cellular biosynthetic process. |
| **steelblueB** | 86 | acute-phase response, cellular protein metabolic process, macromolecule modification, response to wounding, regulation of phosphate/phosphorus metabolic process. |
| **thistle2B** | 51 | blood circulation, circulatory system process, regulation of autophagy, response to drug, cellular response to chemical stimulus. |
| **saddlebrownC** | 81 | DNA biosynthetic process, cellular component assembly, DNA metabolic process, regulation of protein phosphorylation. |

We highlight the “magentaA” module for its GO terms and their implications on the asthma condition. The GO terms expressed in this module are those that are responsible for key ontological pathways, such as cell communication and signaling. As the largest “asthma-unique” module with 274 genes, its differential expression across conditions seems to suggest a larger implication about the disease as a whole; thus, we will select this module for further analysis.

### Topological analysis of the magenta module

We create the hive plots for the magentaA module, shown in Figures 10 and 11, for the correlation networks of the healthy and asthma conditions, respectively.

**Figure 10.**
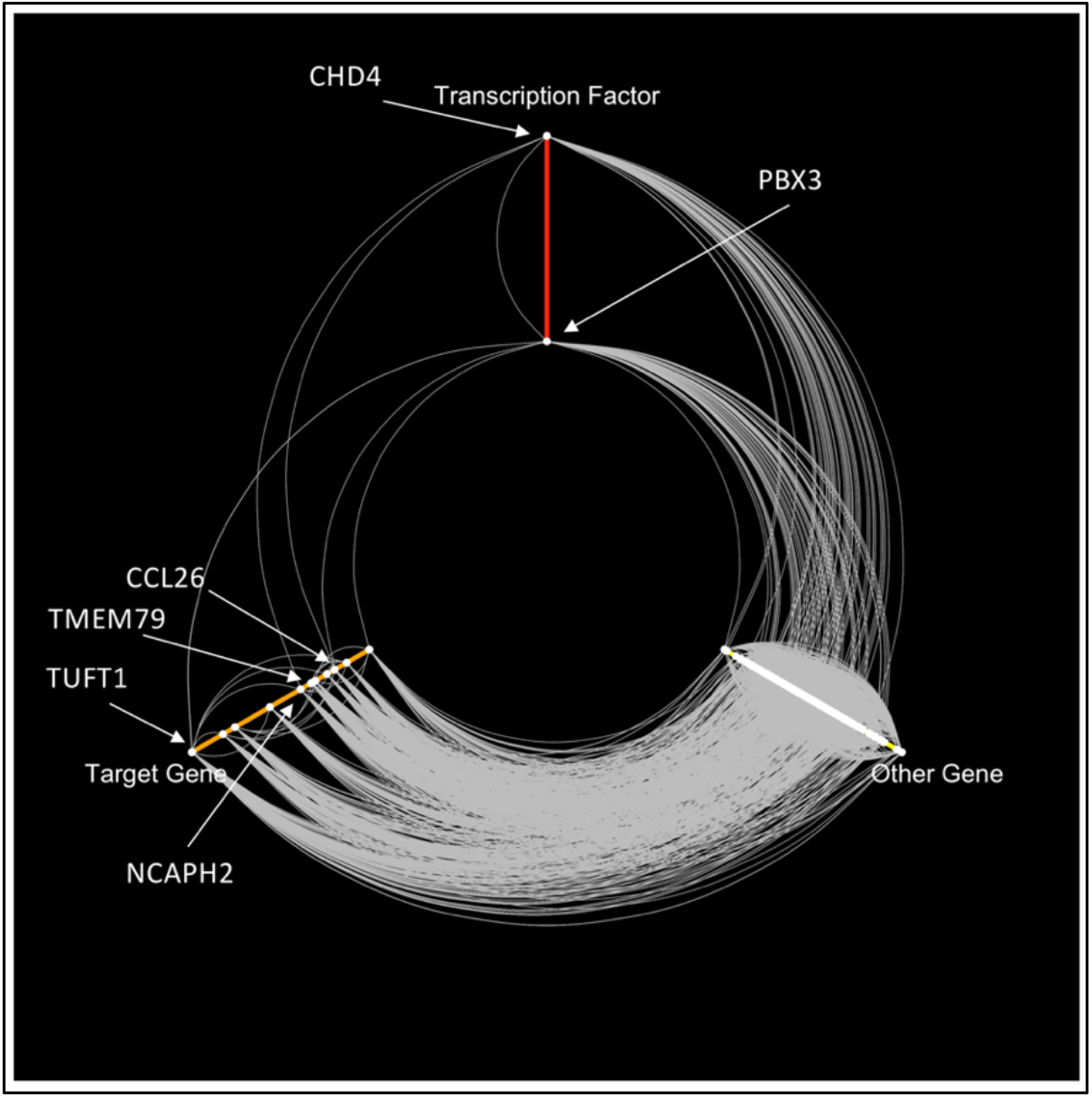
Hive plot of the healthy network of the magenta module. Important interactions include the CHD4-CCL26 and CHD4-TMEM79 connections.

**Figure 11.**
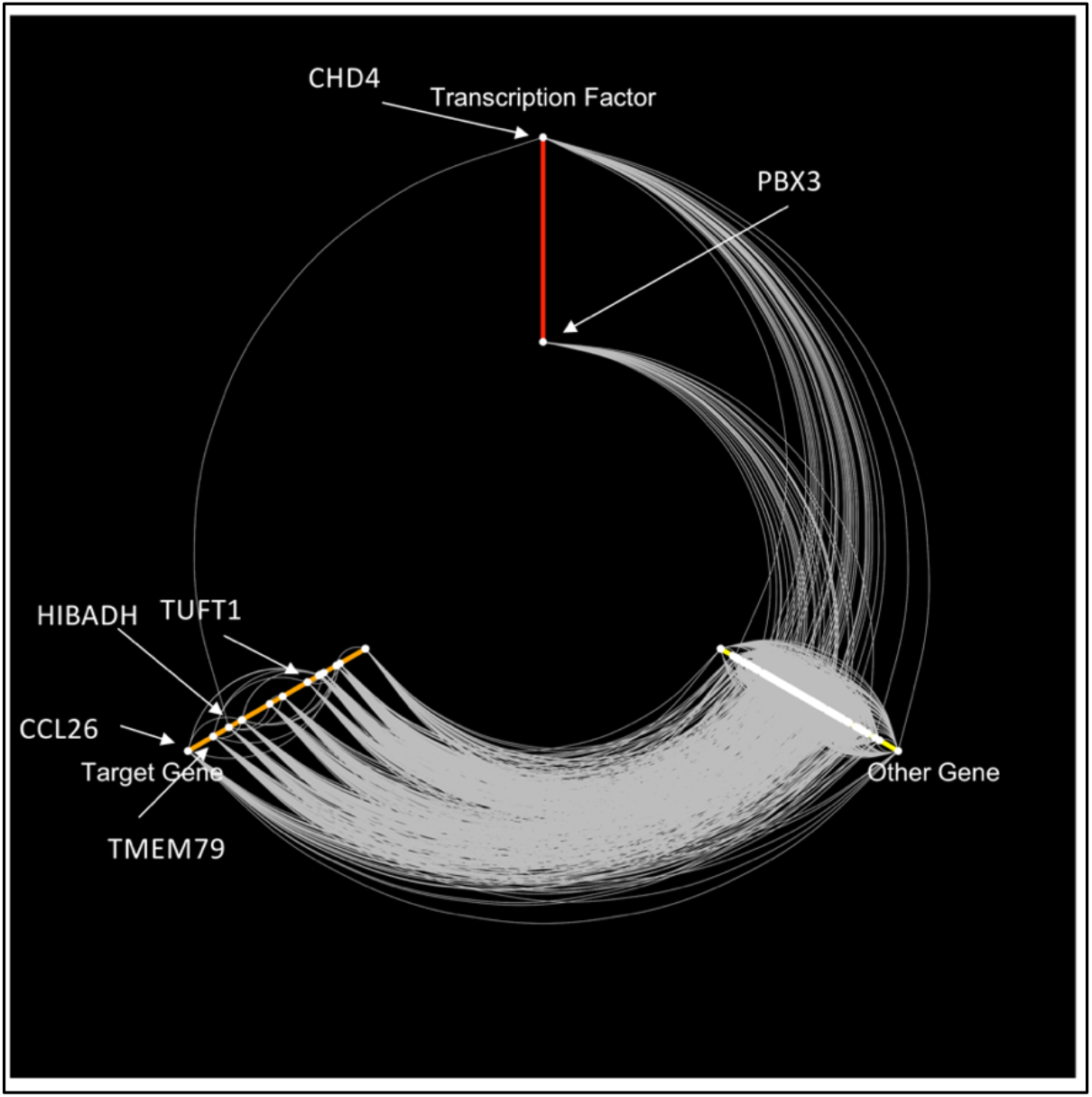
Hive plot of asthma network of the magenta module. Key dysregulations are highlighted with respect to the transcription factors.

## Materials and Methods

### Methodology Overview

In Figure 12, we present a pipeline of the study’s methodology. In short, we use three heterogeneous publicly available datasets (GEO) in conjunction to determine key biomarkers and drug targets in asthma. We first apply the MetaIntegrator framework to determine a set of differentially expressed (DE) genes across all three datasets in asthma. Then, we use the WGCNA framework to determine biological modules that show consistent dysregulation in asthma. We then subset specific networks, based on functional analysis, for topological analysis to determine key interactions and drug targets in asthma.

**Figure 12.**
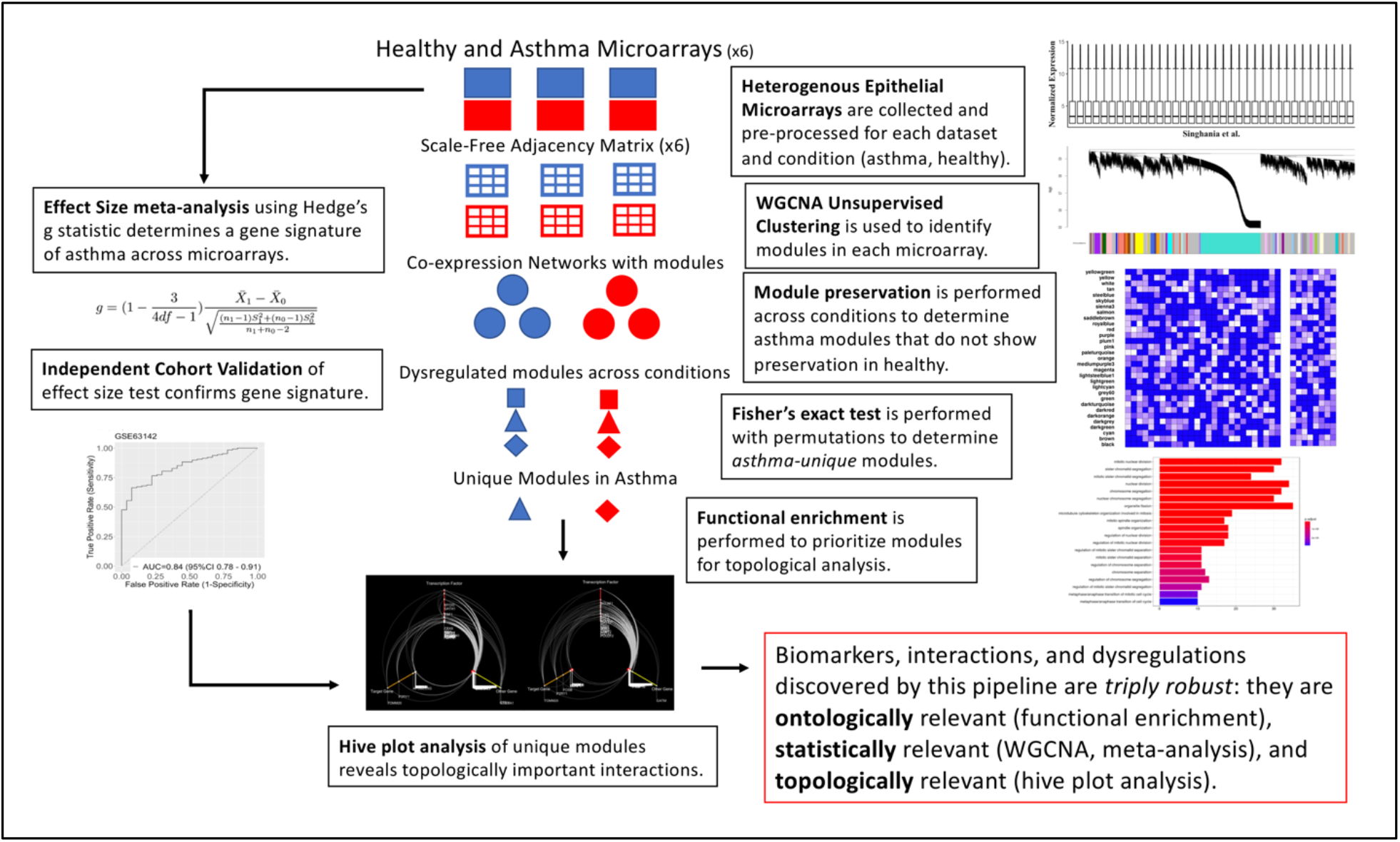
Overview of analysis methodology. Using three microarray matrices, we end up with the comprehensive hive plots that can be analyzed for key interactions and drug targets.

The basis for this pipeline is that it combines an exhaustive series of analyses that test for different forms of significance: statistical, functional, and topological. By combining these forms of analysis, we can be sure that our findings are the rigorous and significant with regards to asthma.

All analysis and figure generation is done by the R software.

### Data pre-processing

Using the three microarray GEO datasets (GSE64913, GSE63142, GSE65204), we use their corresponding GPL files to map the probe readings to gene IDs. In order to establish a bijection between probe readings and gene IDs, we average the readings for those genes which correspond with multiple probes. We then perform quantile normalization across individual studies to convert the raw counts as given by GEO into a data matrix.

### Gene-expression meta-analysis

For our three datasets, we use the MetaIntegrator pipeline to determine a list of differentially expressed genes. We use a mixed models effect size test to combine the relative gene expression values from each of the individual cohorts. Using a two-sided p-value threshold of 0.05, we determine a set of differentially expressed genes in the asthma condition, for both under-expression and overexpression.

### Gene co-expression network reconstruction

The Weighted Gene Co-expression Network Analysis (WGCNA) R package was used to build co-expression networks based on the values in each of our individual cohorts. Through WGCNA, we arrived at six networks, three sets of healthy and asthma networks corresponding to our three studies. Pairwise Pearson’s correlations were used to create the adjacency matrix. Soft-thresholding was used to select powers that yielded scale-free topology.

### Module preservation in networks

The adjacency matrices are used to calculate Topological Overlap Measure (TOM) for each of the networks. We determine biological modules using the dynamic tree cut algorithm at a height of 0.99. Modules are labelled by colors, which have no biological meaning themselves other than as a source of identification. We also identified hub genes in each of the modules.

Module preservation is performed using the WGCNA permutation test, which computes a Zsummary score for each module based on how well this module clusters in a different network. We perform three permutation tests, one for each study, to determine Zsummary scores for each of the modules in asthma. We use the metric outlined by Langfelder et al., which indicates that a module is considered not significantly preserved if it has a Zsummary score less than 10, and strongly preserved otherwise ^32,33^. We choose, across our three comparisons, the modules in asthma that have a Zsummary score less than 10.

### Gene prioritization

We determine the parent GO terms and hub genes associated with the modules that we found to be dysregulated in asthma (in the previous section). For each module, we determine the all the GO terms that are expressed at a 0.001 level of significance. From these terms, we compute the parent GO term based on the gene ontology hierarchy. In each network, we find all such GO terms that are the parent GO term for more than one module. We present these GO terms that correspond with the hub genes of their modules.

### Identification of asthma-unique modules

Our identification of “asthma-unique” modules revolves around the use of the Fisher’s exact test for similarity at a threshold of 0.05: those modules that are highly similar (in terms of their overlapping genes relative to their size) will exhibit smaller p-values and those modules which are completely disjoint will have a p-value of 1.

We perform a pairwise Fisher’s test on all of the *n* dysregulated modules. We then arrive at a *n* × *n* heatmap of all the p-values for the modules. We call a module “asthma-unique” if it has a significant p-value with at least one module from both of the other networks. The rationale behind this algorithm is that a module that is well-preserved in both other studies is likely dysregulated consistently across the disease.

GO term analysis is performed on the “asthma-unique” modules to determine ones which are fruitful for future analysis.

### Topological analysis of key modules

Selected “asthma-unique” modules are subjected to topological analysis via the hive plots. From our original correlation matrices, we develop an aggregate correlation matrix that contains the maximum correlations for the three datasets. We have one aggregate correlation matrix for both healthy and asthma conditions. The gene sets from our selected “asthma-unique” module(s) are used to extract a subnetwork, which we represent using the igraph framework^34^.

Of the genes in these subnetworks, we classify them as one of three types of genes:

- Transcription factors: 114 genes identified by TFDB ^35^
- Differentially expressed genes: genes identified by the MetaIntegrator analysis at p < 0.1
- All other genes

We are particularly interested in the interactions between transcription factors and DE genes, as such an interaction would be doubly relevant to the study of regulatory networks in asthma.

Of our networks, we compute the topological degree of each of the genes, or the number of genes that are connected to each other based on a correlation cutoff. We then use this information to create hive plots with the HiveR package ^36^.

Hive plots are representations of networks along three axes, each representing one of the categories shown above. Genes are plotted categorically on these axes, and their degrees are used to determine quantitatively where they lie on the axes. Genes with high degree are plotted more radially outward than those with lower degrees. Genes that are connected in the graph are then connected. We perform this analysis on the module(s) of interest across healthy and asthma conditions, allowing us to analyze the topology and identify dysregulations.

With these hive plots, we are able to uncover the possible interactions as genes that may serve as drug targets or biomarkers for further exploration.

## Discussion

The magenta module is chosen based on a series of stringent criteria that serve as qualifiers for its high relevance to the asthma disease. The module represents a group of genes that play a common functional role in the genome. Through the permutation test, we identified that its expression is not conserved in healthy condition, which suggests a degree of dysregulation or abnormality. Then, via the Fisher’s test, we found that this module is well-conserved across all three studies as a module that is differentially expressed, or “asthma-unique.” From here, functional analysis using GO terms shows that this module is key for fundamental cell processes, which suggests that its statistical dysregulation has biological implications on asthma as a whole.

Indeed, of the interactions whose differential topologies are highlighted by the hive plots, we find many of the genes to be key biomarkers and genes in established literature. For example, *CHD4* is cited as a key transcription factor that controls Th2 inflammation in asthma ^37^.Because it seems that the *CHD4* complex is a regulator of Th2 inflammation, a known biomarker of asthma, the sparse topology in Figure 11 as compared to Figure 10 seems to confirm that a dysregulation of this nature as also a biomarker of asthma. In this way, the *CHD4* complex may serve as a key drug target for the control of inflammation, which is known to be applicable to medicine as asthma control ^38,39^.

Another area of analysis involves the topological behavior of the *CCL26*, whose behavior in Figures 10 and 11 suggest a bit of nuance to be explored through the hive plots. In the healthy condition, we find the intact *CHD4-CCL26* interaction, which seems to align with an indication of a healthy gene network. In the asthma condition shown in Figure 11, however, this interaction is no longer intact; however, *CCL26* shows a great increase in its degree in the asthma network, make it the DE gene with highest degree. *CCL26* is a gene that has been cited as responsible for recruiting eosinophils, which could explain its connection with *CHD4* as being characteristic of the healthy condition ^38,40^. However, according to Larose *et al*., *CCL26* levels have been shown to increase in the asthma condition, which suggest that its prominence may increase in response to asthma. Thus, it seems that the critical difference in *CCL26* may actually depend on its interaction with *CHD4* rather than simply its over-expression. Future biological analysis should take into account this nuance with regards to *CCL26*: it does serve as a biomarker for asthma, but its expression level and degree are less meaningful than its interactions with transcription factors.

Ultimately, this analysis pipeline is able to, with high levels of confidence, explore the complexity of the CHD4-CCL26 interaction, which seems to have biological implications beyond the simple “over-expression, under-expression” framework that is typical in classic bioinformatics pipelines.

## Data Availability

All data is available on the GEO database and is cited in the paper. Processed forms of these datasets are included as supplemental files.

## Acknowledgements and Funder Statement

We thank the Stanford Institutes of Medicine Summer Research Program (SIMR) for supporting this paper and the Nadeau Lab members for providing feedback on the research.

This study was supported from funding by the Strober Family Fund. The funders had no role in the designing of the research, decision to publish, or authoring of manuscript.

## Supplementary Files

Figure S1. Box plots of quantile-normalized gene expression matrices.

File 1. Zip file containing all six matrices (.csv) separated by study and condition, after aggregation of probe readings, removal of missing values, and quantile normalization across studies.

File 2. List of DE genes as given by MetaIntegrator.

## Notes

### Competing Interest Statement

The authors have declared no competing interest.

### Funding Statement

No external funding was provided for the manuscript.

## References

1. Haidich, A. B. (2010). Meta-analysis in medical research. Hippokratia 14, 29–37 (2010).

2. Loo, S. L. & Wark, P. A. B. Recent advances in understanding and managing asthma [version 1; referees: 2 approved]. F1000Research vol. 5 (2016).

3. Boulet, L. P. & Boulay, M. È. Asthma-related comorbidities. Expert Review of Respiratory Medicine vol. 5 377–393 (2011).

4. Izuhara, K. & Saito, H. Microarray-based Identification of Novel Biomarkers in Asthma. www.jsaweb.jp! (2006) doi:10.2332/allergolint.55.361.

5. Faiz, A. & Burgess, J. K. How Can Microarrays Unlock Asthma? J. Allergy 2012, 1–15 (2012).

6. Carr, T. F. & Bleecker, E. Asthma heterogeneity and severity. World Allergy Organization Journal vol. 9 (2016).

7. Schloss, P. D. identifying and overcoming threats to reproducibility, replicability, robustness, and generalizability in microbiome research. MBio 9, (2018).

8. Haynes, W. A. et al. Empowering multi-cohort gene expression analysis to increase reproducibility. in Pacific Symposium on Biocomputing vol. 0 144–153 (World Scientific Publishing Co. Pte Ltd, 2017).

9. Miłkowski, M., Hensel, W. M. & Hohol, M. Replicability or reproducibility? On the replication crisis in computational neuroscience and sharing only relevant detail. J. Comput. Neurosci. 45, 163–172 (2018).

10. Grützmann, R. et al. Meta-analysis of microarray data on pancreatic cancer defines a set of commonly dysregulated genes. Oncogene vol. 24 5079–5088 (2005).

11. Walsh, C., Hu, P., Batt, J. & Santos, C. Microarray Meta-Analysis and Cross-Platform Normalization: Integrative Genomics for Robust Biomarker Discovery. Microarrays 4, 389–406 (2015).

12. Wang, C., Li, H., Cao, L. & Wang, G. Identification of differentially expressed genes associated with asthma in children based on the bioanalysis of the regulatory network. Mol. Med. Rep. 18, 2153–2163 (2018).

13. Bakhtiarizadeh, M. R., Hosseinpour, B., Shahhoseini, M., Korte, A. & Gifani, P. Weighted gene co-expression network analysis of endometriosis and identification of functional modules associated with its main hallmarks. Front. Genet. 9, (2018).

14. Mao, Z. et al. Transcriptional regulation on the gene expression signature in combined allergic rhinitis and asthma syndrome. Epigenomics 10, 119–131 (2018).

15. García-Campos, M. A., Espinal-Enríquez, J. & Hernández-Lemus, E. Pathway analysis: State of the art. Frontiers in Physiology vol. 6 (2015).

16. Curtis, R. K., Orešič, M. & Vidal-Puig, A. Pathways to the analysis of microarray data. Trends in Biotechnology vol. 23 429–435 (2005).

17. Fehrenbach, H., Wagner, C. & Wegmann, M. Airway remodeling in asthma: what really matters. Cell and Tissue Research vol. 367 551–569 (2017).

18. Hansel, N. N. & Diette, G. B. Gene expression profiling in human asthma. Proceedings of the American Thoracic Society vol. 4 32–36 (2007).

19. Tsai, Y. H., Parker, J. S., Yang, I. V. & Kelada, S. N. P. Meta-analysis of airway epithelium gene expression in asthma. Eur. Respir. J. 51, (2018).

20. Kudo, M., Ishigatsubo, Y. & Aoki, I. Pulm_pdf_L06_PathAsthma. Front. Microbiol. 4, 263 (2013).

21. Pascoe, C. D. et al. Gene expression analysis in asthma using a targeted multiplex array. BMC Pulm. Med. 17, (2017).

22. Park, H. W. et al. Assessment of genetic factor and depression interactions for asthma symptom severity in cohorts of childhood and elderly asthmatics. Exp. Mol. Med. 50, (2018).

23. Rothenberg, M. E. CD44 — a sticky target for asthma. J. Clin. Invest. 111, 1460–1462 (2003).

24. Schori, H., Shechter, R., Shachar, I. & Schwartz, M. Genetic Manipulation of CD74 in Mouse Strains of Different Backgrounds Can Result in Opposite Responses to Central Nervous System Injury. J. Immunol. 178, 163–171 (2007).

25. Borish, L. & Culp, J. A. Asthma: A syndrome composed of heterogeneous diseases. Ann. Allergy, Asthma Immunol. 101, 1–9 (2008).

26. Chen, Y.-A., Tripathi, L. P. & Mizuguchi, K. TargetMine, an Integrated Data Warehouse for Candidate Gene Prioritisation and Target Discovery. PLoS One 6, e17844 (2011).

27. Benjamini, Yoav; Hochberg, Y. Controlling the False Discovery Rate - a Practical and Powerful Approach to Multiple Testing. Journal of the Royal Statistical Society Series B-Methodological 1995.pdf. J. R. Stat. Soc. Ser. B 57, 289–300 (1995).

28. Zhu, J. & Paul, W. E. Peripheral CD4+ T-cell differentiation regulated by networks of cytokines and transcription factors. Immunol. Rev. 238, 247–262 (2010).

29. Albert, R. Scale-free networks in cell biology. Journal of Cell Science vol. 118 4947–4957 (2005).

30. Teschendorff, A. E., Banerji, C. R. S., Severini, S., Kuehn, R. & Sollich, P. Increased signaling entropy in cancer requires the scale-free property of protein interaction networks. Sci. Rep. 5, (2015).

31. Torgerson, D. G. et al. Meta-analysis of genome-wide association studies of asthma in ethnically diverse North American populations. Nat. Genet. 43, 887–892 (2011).

32. Langfelder, P. & Horvath, S. WGCNA: an R package for weighted correlation network analysis. BMC Bioinformatics 9, 559 (2008).

33. Langfelder, P., Luo, R., Oldham, M. C. & Horvath, S. Is my network module preserved and reproducible? PLoS Comput. Biol. 7, (2011).

34. Csárdi, G. & Nepusz, T. The igraph software package for complex network research.

35. Hu, H. et al. AnimalTFDB 3.0: a comprehensive resource for annotation and prediction of animal transcription factors. Nucleic Acids Res. 47, D33–D38 (2019).

36. Krzywinski, M., Birol, I., Jones, S. J. & Marra, M. A. Hive plots--rational approach to visualizing networks. Brief. Bioinform. 13, 627–644 (2012).

37. Hosokawa, H. et al. Functionally distinct Gata3/Chd4 complexes coordinately establish T helper 2 (Th2) cell identity. Proc. Natl. Acad. Sci. U. S. A. 110, 4691–4696 (2013).

38. Larose, M. C. et al. Correlation between CCL26 production by human bronchial epithelial cells and airway eosinophils: Involvement in patients with severe eosinophilic asthma. J. Allergy Clin. Immunol. 136, 904–913 (2015).

39. Diamant, Z. et al. Towards clinically applicable biomarkers for asthma – An EAACI position paper. Allergy (2019) doi:10.1111/all.13806.

40. Silkoff, P. E. et al. Identification of airway mucosal type 2 inflammation by using clinical biomarkers in asthmatic patients. J. Allergy Clin. Immunol. 140, 710–719 (2017).

